# First-in-human autologous oral mucosal epithelial sheet transplantation to prevent anastomotic re-stenosis in congenital esophageal atresia

**DOI:** 10.1101/2021.08.15.21262092

**Authors:** Akihiro Fujino, Yasushi Fuchimoto, Yoshiyuki Baba, Nobutaka Isogawa, Takanori Iwata, Katsuhiro Arai, Makoto Abe, Nobuo Kanai, Ryo Takagi, Masanori Maeda, Akihiro Umezawa

## Abstract

**Background:** Congenital esophageal atresia postoperative anastomotic stricture occurs in 30-50% of cases. Patients with severe dysphagia are treated with endoscopic balloon dilatation (EBD) and/or local injection of steroids, but many patients continue to experience frequent stricture. In this study, we investigated the transplantation of autologous oral mucosa-derived cell sheets (epithelial cell sheets) as a prophylactic treatment for congenital esophageal atresia postoperative anastomotic stricture.

**Methods:** Epithelial cell sheets were fabricated from a patient’s oral epithelial tissue, and their safety was confirmed by quality control tests. The epithelial cell sheets were transported under controlled conditions from the fabrication facility to the transplantation facility and successfully transplanted onto the lacerations caused by EBD using a newly developed transplantation device for pediatric patients. The safety of the transplantation was confirmed by follow-up examinations over 48 weeks.

**Results:** The number of EBDs required after transplantation and the number of days between EDBs were recorded. Before transplantation, EBDs were performed approximately every two weeks, whereas after transplantation, the interval was extended to a maximum of four weeks. The patient was also aware of a reduction in dysphagia.

**Conclusions:** This study suggests that cell sheet transplantation might be effective in preventing anastomotic stricture after surgery for congenital esophageal atresia. We chose this very severe case for the first clinical study in humans. Future studies are needed to identify cases in which cell sheet transplantation is most effective and to determine the appropriate timeframes for transplantation.

## Background

Anastomotic stricture after surgery for congenital esophageal atresia is reported to occur in 30-50% of cases [1–3]. The main symptom of anastomotic stricture is dysphagia. Some patients with anastomotic stricture are unable to swallow saliva and require continuous saliva aspiration. Patients with feeding difficulties, such as choking on food, are treated with endoscopic balloon dilation (EBD) and/or local injections of steroids, but many patients have repeated stenosis even after frequent EBD and/or local injections of steroids. In an analysis of 884 cases of esophageal atresia, anastomotic stricture was reported to occur in 40% of all cases, and of these, anastomotic dilation was ineffective and resection of the stricture was performed in 2-7% [1]. Sarhal et al. reported that 23 of 62 patients (37%) had anastomotic stenosis and required dilatation at an age of 149 days (mean value, the range was 30-600 days), and stenosis was relieved by 3.2 dilations (mean value, the range was 1-7 dilations), but 3 patients experienced severe persistent dysphagia [2]. Antoniou et al. reported that a total of 165 dilations, ranging from 1 to 9 dilations per patient (median value 2.79 dilations), was effective in 59 of 167 patients, while it was ineffective in 12 patients (20.3%) [3]. Because balloon dilation under fluoroscopy can cause a great deal of physical and mental distress to the patient, EBD often requires general anesthesia in pediatric patients. For these reasons, patients often become reluctant to consume food and are unable to enjoy it. This leads to malnutrition and poor growth. Repeated anastomotic stricture after esophageal atresia increases the physical and mental burden on the patient and significantly impairs quality of life. Therefore, in recent years, the resection of the stricture and reanastomosis, or implantation of an absorbable stent have been recommended for patients overseas who suffer restenosis even after four EBDs [1]. However, multiple surgeries may be life-threatening, and the possibility of restenosis is high. Absorbable stenting is also associated with restenosis after stent resorption, and there are no ideal treatments for anastomotic stenosis [4].

Recently, transplantation of autologous oral mucosa-derived cell sheets (hereinafter referred to as “epithelial cell sheets”) has been shown to be very useful for esophageal stricture that almost inevitably occurs after extensive endoscopic submucosal dissection (ESD) for superficial esophageal squamous cell carcinoma (ESCC) in adults [5]. Okano et al., at the Institute of Advanced Biomedical Engineering and Science (ABMES), Tokyo Women’s Medical University, proposed “cell sheet engineering”, in which isolated cells are cultured on special inserts called “temperature-responsive cell culture inserts” and then processed into sheets for use by the properties of the somatic stem cells. This non-invasive method allows cells to be peeled off and collected as a cell sheet by simply lowering the temperature without the use of destructive enzymes such as trypsin, thus enabling sutureless cell engraftment while retaining the integrity of extracellular matrix proteins, such as cell adhesion factors. In addition, since the cells are processed into a sheet that can be transplanted to a target site, the applicable range of transplantation sites can be greatly expanded.

ABMES developed a successful method for prevention of esophageal stricture after ESD using tissue-engineered cell sheets. Epithelial cell sheets were endoscopically transplanted onto esophageal ulcer surfaces after ESD, which showed promising clinical results in preventing esophageal stricture [5,6]. From 2008 to 2010, ten adults with esophageal stricture after extensive ESD for ESCC were treated with this therapy, and esophageal stricture was prevented in nine out of ten patients. From 2013 to 2014, ten adults with esophageal stricture after extensive ESD for ESCC were treated with this therapy at Nagasaki University Hospital [7]. In this case, oral mucosal tissue collected at Nagasaki University Hospital was transported to the cell processing facility (CPF) at Tokyo Women’s Medical University, where epithelial cell sheets were produced, and then returned to Nagasaki University Hospital for transplantation. Of the ten cases of ESCC with a circumference of 80% or more, where stricture is generally inevitable after ESD, positive results were obtained; esophageal stricture was prevented in six cases. Patients were followed up with physical and clinical examinations for 48 weeks after transplantation, and the safety of the cell sheet transplantation was confirmed.

Based on the results of previous clinical studies of autologous cell sheet transplantation after ESD in adult patients with ESCC, this study was designed to investigate autologous cell sheet transplantation as a prophylactic treatment for postoperative anastomotic restenosis of congenital esophageal atresia. Prior to clinical trials in humans, preclinical studies were performed in a porcine model [8]. Miniature pigs with simulated esophageal stricture were divided into two groups: one received conventional EBD alone, and the other received both EBD and epithelial cell sheet transplantation, and the degree of restenosis after two weeks was compared. The results suggested that cell sheet transplantation was effective in reducing restenosis. After confirming the safety and efficacy of cell sheet transplantation in this porcine model, we performed a clinical trial on a human subject. This study also determined that the autologous oral cavity mucosa-derived cell sheets could be successfully transported from the fabrication facility to the hospital where the transplantation was performed on the patient.

## Methods

### Subject selection criteria

The subjects of this study were selected from patients with anastomotic stricture in postoperative congenital esophageal atresia who still had a stricture after at least five EBDs. Patients between the ages of 1 and 18 years were included in this study. Patients suffering from other serious diseases (including heart, kidney, and liver diseases) or severe anemia (hemoglobin level less than 10.0 g/dl) were excluded from this study. This clinical study was conducted on one patient who met these conditions.

### Fabrication of oral mucosal epithelial cell sheets

In accordance with previous studies, cultured autologous oral mucosal epithelial cell sheets were fabricated by using buccal mucosal tissue and serum derived from the patient at the CPF of Tokyo Women’s Medical University [7,9]. Procedures for fabrication of the epithelial cell sheets are described in standard operating procedures (SOP), and the procedures were carried out according to the SOPs. In brief, buccal mucosal tissue was harvested from the oral cavity of the patient at the National Center for Child Health and Development (NCCHD) and put into a 50 mL centrifuge tube with Dulbecco’s modified Eagle’s medium (DMEM) (Sigma-Aldrich, MO, USA) supplemented with 57.1 µg/mL ampicillin and 28.6 µg/mL sulbactam (Unasin-S, Pfizer, NY, USA), 100 µg/mL streptomycin (Meiji Seika Pharma, Tokyo, Japan), and 1.0 µg/mL amphotericin B (Fungizone, Bristol-Myers Squibb, NY, USA). Blood derived from the patient was also collected into 50 mL centrifuge tubes. The centrifuge tubes with the mucosal tissue and the blood were transported to the CPF at 4°C and room temperature, respectively. Autologous serum of the patient was prepared by the conventional method; centrifugation two times at 1,830*xg*, and used as a supplement for culture medium. The oral mucosal cells were prepared by treatment with 1,000 U/mL Dispase I (Godo Shusei, Chiba, Japan) for two hours at 37°C and 2.5 mg/mL trypsin and 380 µg/mL ethylenediamine tetraacetic acid tetrasodium (EDTA) salt dihydrate (Thermo Fisher Scientific, MA, USA) for 20 min at 37°C. The disaggregated oral mucosal cells were suspended in a keratinocyte culture medium (KCM) composed of a basal medium consisting of three parts of DMEM and one part of nutrient mixture F-12 Ham (Sigma-Aldrich) supplemented with 40 µg/mL gentamicin (Gentacin, Takata Pharmaceutical, Saitama, Japan), 0.25 µg/mL amphotericin B (Fungizone, Bristol-Myers Squibb, NY, USA), 5% autologous serum, 5 µg/mL insulin (Humulin; Eli Lilly, IN, USA), 10 ng/mL human recombinant epidermal growth factor (Higeta-Syoyu, Chiba, Japan), 1 ng/mL cholera toxin (List Biological Laboratories, CA, USA), 2 nmol/L triiodothyronine (Fujifilm Wako Pure Chemicals), and 0.4 µg/mL hydrocortisone (Saxizon, Teva Takeda Pharma, Aichi, Japan). The epithelial cells were seeded onto temperature-responsive cell culture inserts (CellSeed, Tokyo, Japan) at a density of 7.5 ×10^4^ cells/cm^2^ and cultured in KCM at 37°C under 5% CO_2_ in the cell culture incubator. Medium replacement was implemented according to the SOP, and the supernatant fractions were used for quality control test. After passing quality control, the epithelial cells were transported to the National Center for Child Health and Development at 37°C. Before transplantation of the epithelial cell sheets onto the esophageal wound, the epithelial cells were harvested as a cell sheet by reducing the temperature from 37°C to room temperature for retrieval by surgical forceps.

### Quality control tests

In order to define the decision criteria for transplantation of cultured autologous oral mucosal epithelial cell sheets, a product standard code was prepared before starting this clinical study. Quality control tests were conducted in accordance with the SOPs for certificating the criteria. Culture media collected after media replacement were subjected to sterility tests consisting of (1) cultivation for aerobic and anaerobic bacteria and fungi, (2) endotoxin detection by a limulus amoebocyte lysate assay, and (3) mycoplasma testing by cultivation and detection of DNA derived from *mycoplasma pneumoniae* by loop-mediated isothermal amplification. These quality control tests were outsourced to SRL, Inc. (Tokyo, Japan). Quantification for cellular density, viability, and epithelial cell percentage of the cultured autologous oral mucosal epithelial cell sheets were implemented using similar protocols to a previous study [10]. In brief, the epithelial cell sheet was harvested from the temperature-responsive culture insert and treated with trypsin EDTA for 20 min at 37°C. The disaggregated oral mucosal cells were suspended in KCM and counted with a hemocytometer to calculate cellular density of the epithelial cell sheet. Quantification of cellular viability was determined by the trypan blue dye exclusion assay, and flow cytometry was employed to determine the percentage of oral mucosal epithelial cell in the cell sheet. The BD Cytofix/Cytoperm kit (BD Biosciences, NJ, USA) and Anti-Cytokeratin/Keratin Type II, FITC (American Research Products, MA, USA) were used in accordance with manufacturer’s protocols. Flow cytometry analysis was performed with Gallios Software Ver.1.2 and Kaluza 1.3 ver1.0 (Beckman Coulter, CA, USA).

### Endoscopic balloon dilatation (EBD)

Dilatation was performed using an esophageal balloon dilatation catheter (CRE Fixed Wire, Boston Scientific Corporation, Natick, Massachusetts, USA). The endoscope (XQ240, Olympus, Tokyo, Japan) was inserted to the location of the stricture and the guidewire was passed through the biopsy channel of the endoscope. Based on the estimated situation of the stricture (location, contour, and diameter), a balloon size of 20 mm was selected. The endoscope was removed with the wire remaining, and the balloon catheter was placed in the center of the stricture under fluoroscopy. The balloon was injected with radiopaque contrast medium to 6 atm for 180 seconds. This was repeated three times.

### Pediatric cell sheet transplantation device

A novel device for endoscopic cell sheet transplantation was designed. The device was composed of two parts, a cell sheet carrier and an air tube, as shown in Figure 3. The carrier was comprised of a body in the shape of a boot, and a medical grade latex balloon sheet which was glued to the body with a biocompatible cyanoacrylate instant adhesive. A female screw as a connector was installed at the proximal end of the body to connect with the air tube. The latex balloon sheet was originally fabricated using the material and production method for ultrasound endoscope balloons. The air tube consisted of a polyurethane tube, stainless male screw connector and proximal hub, attached to a syringe. Since steel wires were braided inside the tube wall, the tube could transfer linear rotation torque from the proximal end to the distal end. The carrier was connected to the air tube in an airtight manner, and was passed through a biopsy channel (> 2.0 mm) of an endoscope. Through the air tube, an operator could supply air into the carrier to expand the balloon and use suction to remove air from the carrier with a syringe. A cell sheet was loaded onto the balloon of the carrier by water surface tension. Then, the cell sheet was placed inside the carrier by air suction, which led the balloon with the cell sheet to adhere on the inner wall of the carrier. Then, the cell sheet could be reliably delivered to the transplantation site in the esophagus through an overtube. After delivery and alignment of the cell sheet within the site, the cell sheet could be attached to the site by balloon inflation to achieve transplantation.

### Endoscopic transplantation of the cell sheet

Two weeks after EBD, cell sheet transplantation was performed using the specialized endoscopic device. Epithelial cell sheets were collected from the culture inserts with the temperature lowered to 20°C and attached to the esophageal laceration created by EBD. Three epithelial cell sheets were used for transplantation.

### Follow-up examination

Follow-up examinations were conducted for about 48 weeks after transplantation of the cell sheets, including periodic endoscopy and clinical examinations. Eight evaluations were performed during follow-up: (1) measurement of vital signs, (2) determination of difficulty in swallowing and whether food stuck in the patient’s throat, (3) blood tests, (4) X-rays, (5) presence of stenosis by endoscopic examination, (6) sheet fixing and epithelialization by endoscopic examination, (7) presence of stenosis by esophagography, and (8) adverse events. The timing of each test is summarized in Supplemental Table 1. If symptoms such as difficulty in swallowing occurred, endoscopic examination or esophagography were performed as needed.

## Results

### Fabrication of epithelial cell sheets

Autologous epithelial cell sheets were prepared from oral epithelial tissue of a patient with congenital esophageal atresia (Figure 1). Mucosal epithelial tissue of the patient’s left cheek was cut with a scalpel, processed, and transported to a biosafety cabinet at the CPF (Figure 2A and 2B). After treatment with Dispase I and trypsin-EDTA, cell viability and concentration were measured by a trypan blue dye exclusion assay and a hemocytemeter (Figure 2C). Viable oral mucosal cells were successfully prepared from the tissue and seeded at a density of 7.50 ×10^4^ cells/cm^2^ onto four temperature-responsive cell culture inserts (Table 1). Adhesion and proliferation of the epithelial cells were observed on the culture surfaces in KCM supplemented with autologous serum (Figure 2D and 2G). Quality control tests of culture supernatants indicated that there was no contamination with bacteria, fungi, and mycoplasma in the epithelial cell sheets, and lower endotoxin concentrations than the established criterion (Table 2). The oral mucosal epithelial cell sheets were harvested from the culture insert and used for quality control tests including cell count, dye exclusion assay, and flow cytometry (Figure 2E and 2F, Table 2).

**Figure 1.**
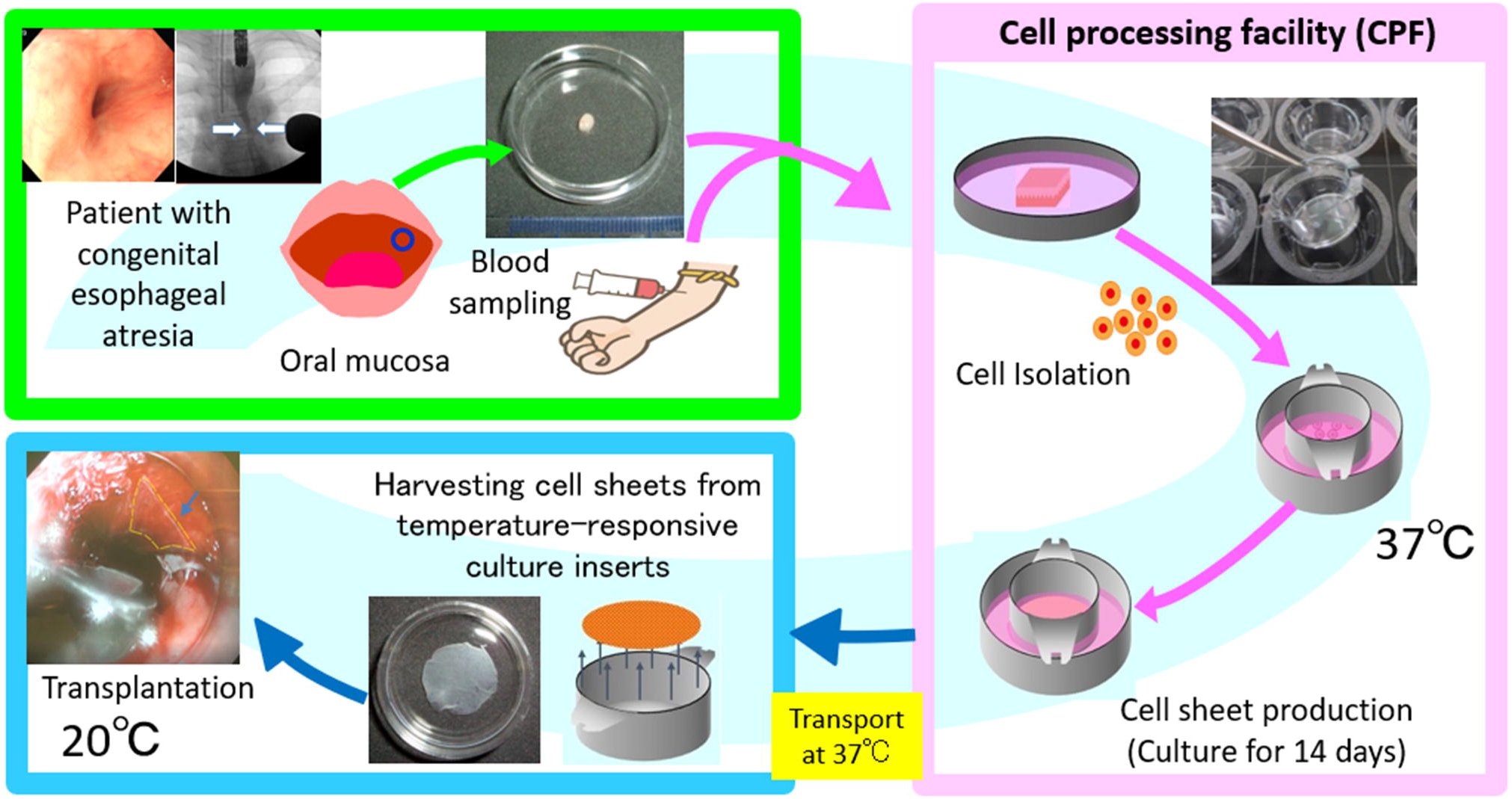
Procedure for transplantation of an autologous cultivated oral mucosal epithelial cell sheet to the anastomotic stenosis in postoperative congenital esophageal atresia. The patient’s oral mucosal tissue and blood were collected at the National Center for Child Health and Development (NCCHD) and transported to the CPF of Tokyo Women’s Medical University while maintaining them at low temperature. The harvested oral mucosal epithelial cells were isolated and seeded onto temperature responsive cell culture inserts. The cells were cultured for 14 days to produce cell sheets. In-process quality control tests were performed at each stage of the cell sheet fabrication process. In addition, a pre-shipment test (quality assessment) was performed. The products that passed the tests were transported to the NCCHD, maintaining the temperature around 37°C. The cell sheets were then transplanted into the patient.

**Figure 2.**
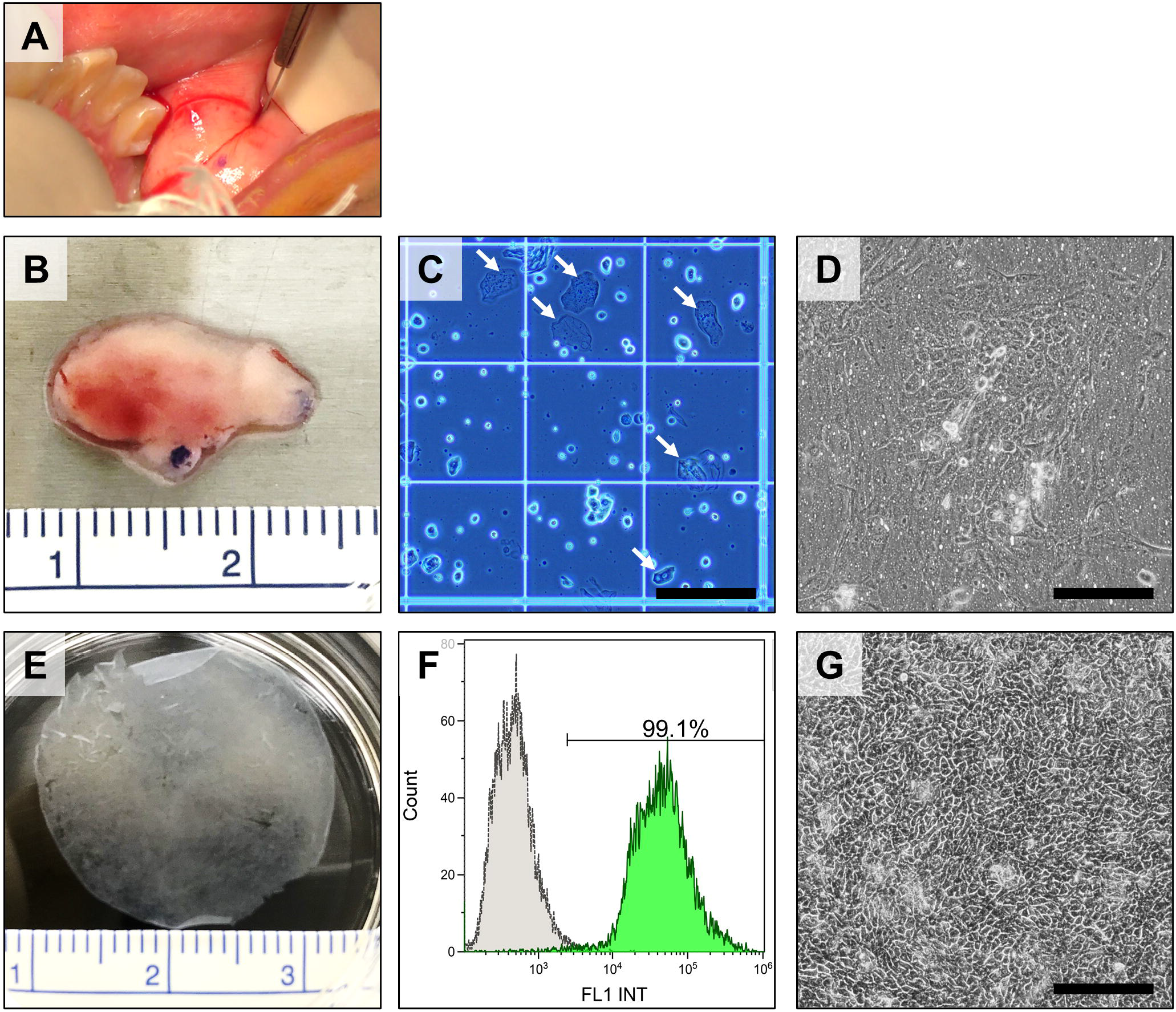
Fabrication and quality control tests for cultured autologous oral mucosal epithelial cell sheets. **(A)** Collection of oral mucosal tissue from the patient. **(B)** Buccal mucosal tissue harvested from the oral cavity of this patient. The superficial area of the mucosal tissue was approximately 0.8 cm^2^. **(C)** Cellular morphology of oral mucosal cells before seeding onto temperature responsive culture inserts. Cells marked with white arrows were terminally differentiated epithelial cells which were not counted as viable cells for determining seeding density of the cells. Scale bar is 200 µm. **(D)** Cellular morphology of the oral mucosal cells 8 days after seeding. Scale bar is 200 µm. **(E)** Cultured autologous oral mucosal epithelial cell sheet for quality control testing for quantification of cellular density, viability, and percentage of epithelial cells. **(F)** Histogram of flow cytometry results for determining percentage of epithelial cells in the cell sheet by detection of cytokeratin-positive cells. Percentage of cytokeratin-positive cells determined to be epithelial cells was 99.1% in the epithelial cell sheet. **(G)** Cellular morphology of the oral mucosal cells before transportation to the hospital. Scale bar is 200 µm.

**Table 1.** Preparation of oral mucosal epithelial cell sheets

|  | Results |
| --- | --- |
| Size of mucosal tissue | 0.818 cm<sup>2</sup> |
| Total cells/tissue | 1.29 x10<sup>6</sup> cells |
| cells/cm<sup>2</sup> of tissue | 1.57 x10<sup>6</sup> cells/cm<sup>2</sup> |
| Viability | 91.0% |
| Seeding density | 7.50 x10<sup>4</sup> cells/cm<sup>2</sup> |
| Number of cell sheets | 4 |

**Table 2.** Quality control tests for oral mucosal epithelial cell sheets

| Table 2. Quality control tests for oral mucosal epithelial cell sheets |  |  |
| --- | --- | --- |
|  | Criteria | Results |
| Sterility tests |  |  |
| aerobic bacteria | (-) | (-) |
| anaerobic bacteria | (-) | (-) |
| fungi | (-) | (-) |
| Endotoxin test | < 1.0 EU/mL | 0.260 EU/mL |
| Mycoplasma tests |  |  |
| cultivation | (-) | (-) |
| DNA of *M. pneumoniae* | (-) | (-) |
| Quality of the sheet |  |  |
| Total cells | $>1.0 \times 10^5$ cells/sheet | $2.14 \times 10^5$ cells/sheet |
| Cellular density | $>2.38 \times 10^4$ cell/cm<sup>2</sup> | $5.09 \times 10^4$ cell/cm<sup>2</sup> |
| Viability | >70% | 98.1% |
| Percentage of epithelial cells | >70% | 99.1% |

### Transplantation of epithelial cell sheets using a newly developed device

After confirming the results of the quality control tests, the epithelial cell sheets were transported at 37°C from the CPF of Tokyo Women’s Medical University to the National Center for Child Health and Development (Figure 1). The culture inserts were brought to room temperature, and the epithelial cell sheets that had detached from the inserts were collected. A special device for endoscopic transplantation of cell sheets for pediatric use was newly developed for this clinical study (Figure 3A-C). Using this device, epithelial cell sheets were successfully transplanted onto the wounds of esophageal mucosa caused by EBD (Figure 4A-G). Epithelial cell sheets were placed on the balloon at the tip of the device one at a time, and the cell sheets were attached to the laceration by inflating the balloon (Figure 4E-G). Three epithelial cell sheets were transplanted.

**Figure 3.**
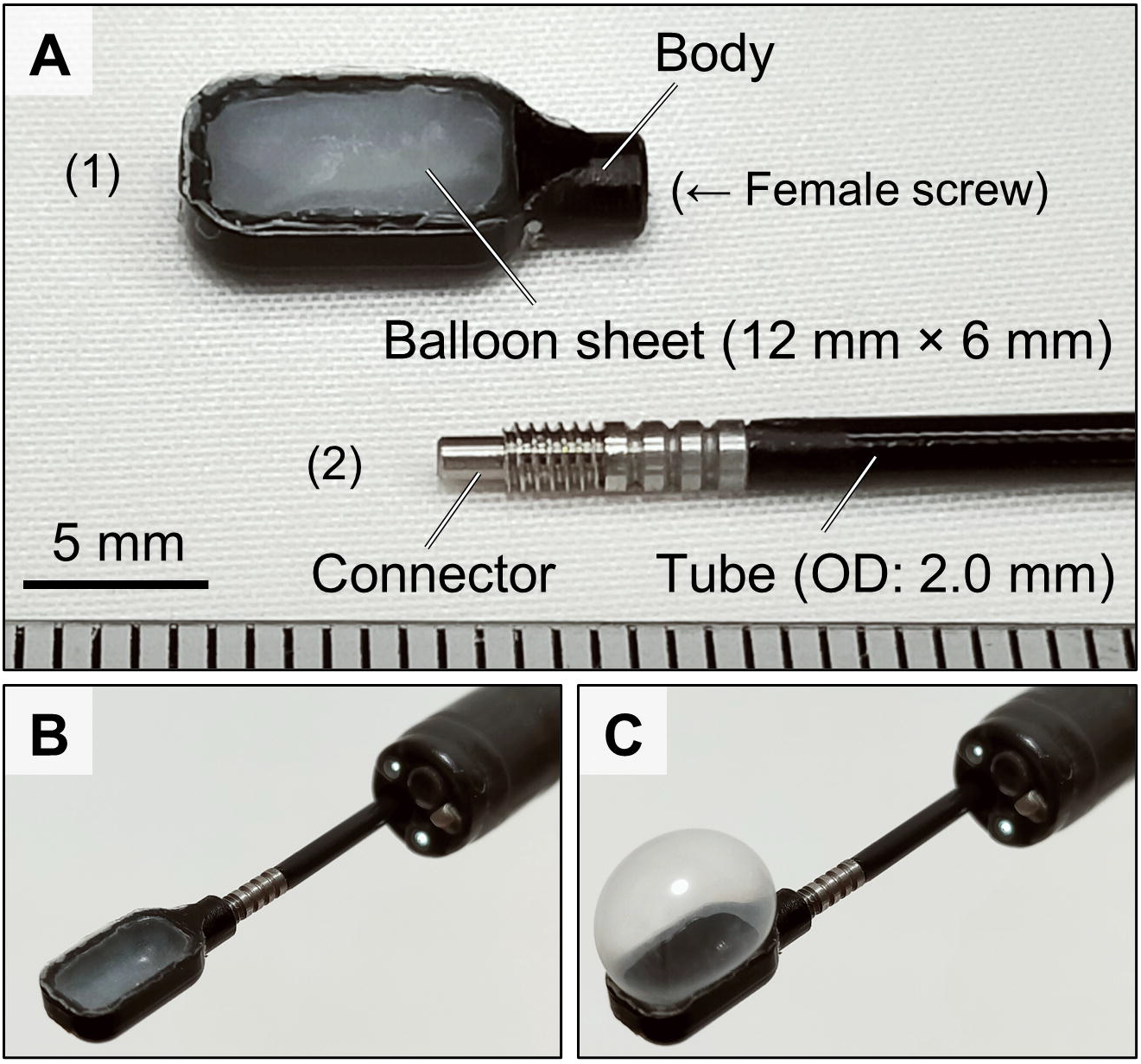
The pediatric cell sheet transplantation device. **(A)** Diagram of the main components. (1) The carrier consisted of a plastic body and latex balloon (0.13 mm thick), which were glued together. (2) The air tube consisted of a steel wire and polyurethane tube with an outer diameter (OD) of 2.0 mm, male screw connector and hub. The air tube was connected to the carrier throughout the endoscopic cannel. (**B**) Delivery mode. A cell sheet was placed inside of the device by suction because the balloon with the cell sheet adhered to the inside wall of the device. By maintaining suction, the cell sheet could easily be delivered to the transplantation site in the esophagus. (**C**) Transplantation mode. The cell sheet is expanded and attached to a transplantation site by balloon inflation.

**Figure 4.**
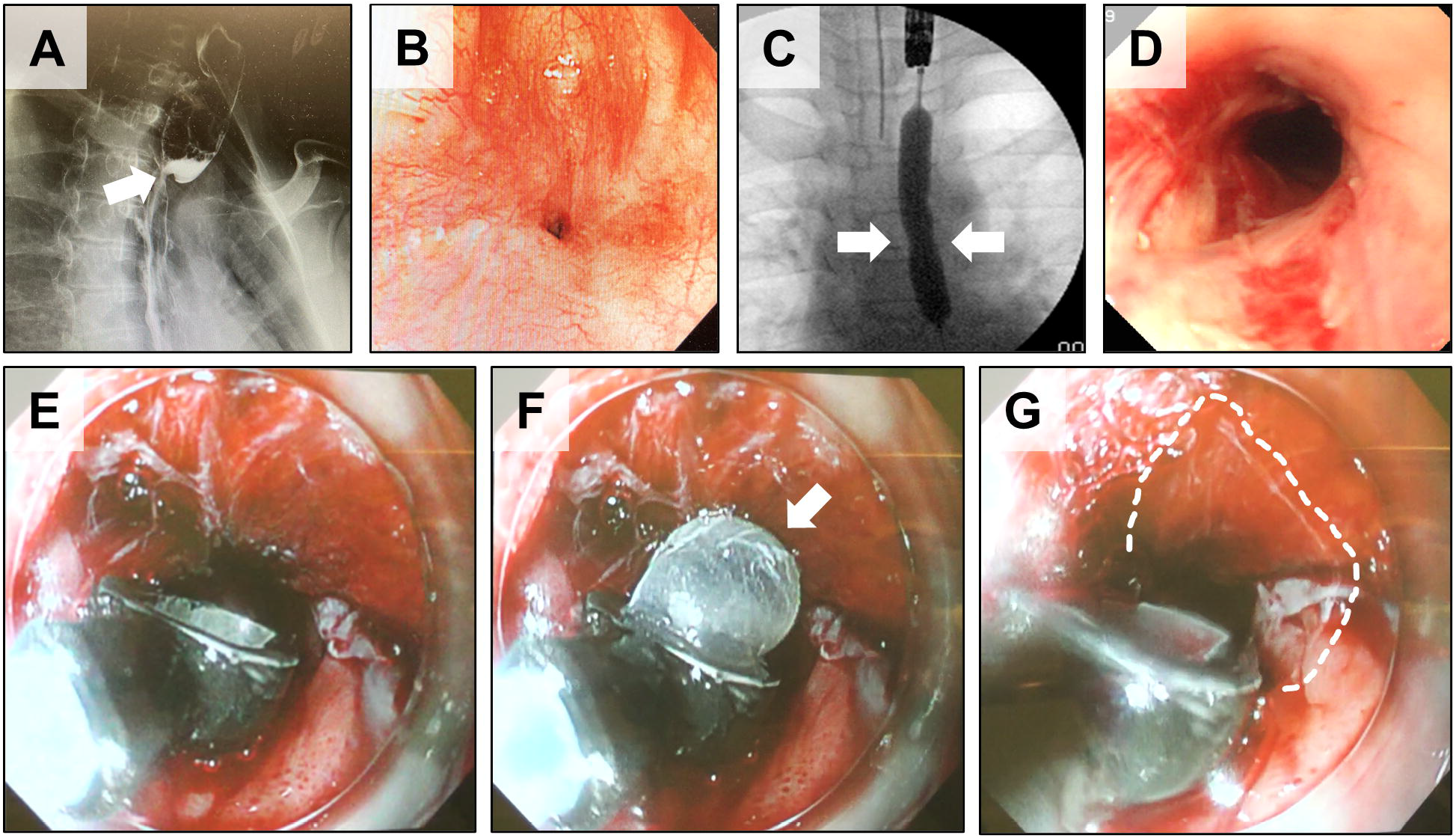
Transplantation of epithelial cell sheets onto anastomotic stenosis using a dedicated device. **(A)** Contrast study findings of the patient’s esophagus before EBD. The area indicated by the arrows is anastomotic stenosis. **(B)** Endoscopic image of the anastomotic stenosis before EBD. **(C)** Contrast study findings of the esophagus during EBD. The area indicated by the arrows was thought to be anastomotic stenosis, which could not be fully expanded due to scar contracture. **(D)** Endoscopic image of the esophagus dilated by balloon and lacerations therein. **(E-G)** Transplantation of cell sheets onto laceration areas caused by EBD using a dedicated device. A cell sheet was set on the surface of the balloon (arrowhead) at the tip of the device (F), and by expanding this balloon, the cell sheet was attached to the lacerated area on the esophageal mucosa. The area surrounded by the white dashed line shows the transplanted cell sheet (G).

### Effect of epithelial cell sheet transplantation to the anastomotic stenosis

The number of days from transplantation of the epithelial cell sheet to the next required EBD was recorded (Figure 5). EBDs were performed approximately every two weeks when the patient was treated only with EBD, whereas after transplantation, the interval was extended to a maximum of four weeks. The total number of EBDs that the patient received after cell sheet implantation was 19. For the first 10 of these, the intervals between EBDs were longer than before transplantation. In addition, the patient felt that the degree of the difficulty in swallowing was alleviated and the instances of food getting stuck were reduced after cell sheet transplantation.

**Figure 5.**
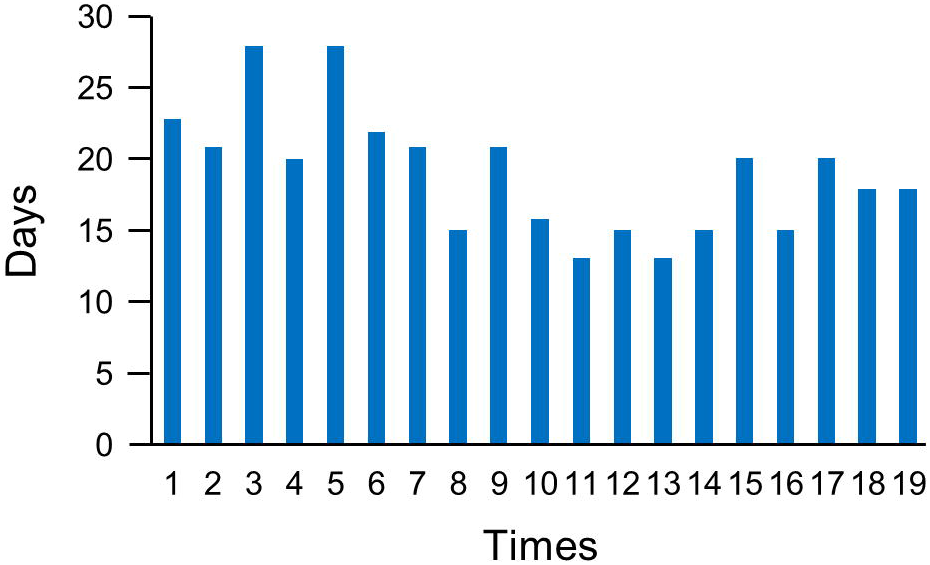
Number of days between EBDs after cell sheet transplantation. Graph of the number of days from cell sheet transplantation to the next EBD, and between procedures. The horizontal axis shows the number of times EBD was performed, and the vertical axis showed the number of days between EBDs.

### Confirmation of safety

Follow-up examinations were performed for about 48 weeks after cell sheet transplantation. The follow-up examination included regular endoscopic and clinical examinations (Supplemental Table 1). All tests showed no abnormalities, confirming the safety of the transplantation.

## Discussion

Epithelial cell sheet transplantation resulted in an increase in the intervals between EBDs. In this case, the criterion for the requirement for EBD was the inability to swallow saliva. Before cell sheet transplantation, the patient’s stenosis became worse about one week after EBD, and the patient was unable to swallow saliva about two weeks after EBD. In other words, the patient needed EBD every two weeks or so. This patient has a more severe case compared to other patients we have seen with the same disease. In contrast, the interval between EBDs was extended to a maximum of four weeks after cell sheet transplantation. In addition, the patient felt that the difficulty associated with swallowing and the degree of food obstruction were alleviated after transplantation. These observations suggest that cell sheet transplantation had an effect. On the other hand, the intervals between EBDs became shorter and eventually reached the same frequency as before transplantation about seven months after transplantation. These results were then evaluated in the context of a case of cell sheet transplantation for adult ESCC.

Histopathological examination of the resected refractory anastomotic stenosis showed tears in the muscularis propria and significant scar tissue in the submucosa [4]. Even if the epithelial cell sheets are transplanted after the mucosa and scar tissue are cut open by EBD, the immediate reforming of the scar tissue will eventually lead to restenosis. However, in this case, the intervals between EBDs were longer for about seven months after transplantation. This suggests that the transplanted cell sheets may have formed a mucosal epithelium that delayed the adhesion and formation of submucosal scar tissue, thereby reducing the degree of scar contracture.

On the other hand, after seven months post-transplantation, the intervals between EBDs became shorter. This is presumably due to the fact that repeated EBD causes progressive fibrosis of the esophagus, allowing scar tissue to re-form at a normal rate and restore the frequency of restenosis. The treatment of congenital esophageal atresia involves wounding of the entire circumference and all layers of the esophagus during the initial resection and anastomosis. In addition, repeated destruction of esophageal tissue by subsequent EBD may promote scar formation and exacerbate anastomotic stenosis. In contrast, in the treatment of adult ESCC with ESD, the mucosa and submucosa of the esophagus are removed, but the muscularis propria and adventitia are retained. This is the main difference between the two treatments. Postoperative anastomoses in patients with congenital esophageal atresia are presumed to have more severe fibrosis than ESD sites for adult ESCC. Therefore, it is assumed that the effect of cell sheet transplantation in this case was not as dramatic as that in ESCC patients.

When anastomotic stricture occurs after esophageal atresia surgery, EBD is recommended as soon as possible to reduce the scar formation at the anastomosis, except during the first month postoperatively, when the risk of perforation is high [3,11]. In addition, it was reported that transplanted autologous skeletal stem cell sheets are effective in myocardial regeneration by releasing cytokines involved in the repair of normal myocardial tissue (this is called the cytokine paracrine effect) [12,13]. Furthermore, in regenerative therapy of articular cartilage, it was reported that if the superficial to shallow layers of cartilage can be repaired and regenerated using cell sheets, and the deeper layers will spontaneously repair themselves later [14]. Based on these reports, in cases where balloon dilation is expected to be necessary more than a certain number of times, early cell sheet transplantation may prevent esophageal stricture severe enough to cause dysphagia by promoting repair and regeneration with retention of native tissue rather than scar tissue. Therefore, future studies are needed to determine the appropriate conditions and timing for esophageal sheet transplantation.

## Conclusions

Usually, when refractory anastomotic stenosis worsens and becomes difficult to treat with EBD, it is necessary to resect the stenosis and connect the non-scarred (normal) portion of the esophagus. However, since the esophagus is considerably stretched and connected in the first anastomosis, suture failure often occurs in the second anastomosis, or there is not enough esophageal length to anastomose an appropriate portion. Therefore, a method to prevent restenosis other than resection and anastomosis is desirable. In particular, this patient refused to have a second resection surgery, so non-surgical treatment options had to be considered. Some patients with esophageal stricture are able to eat through their mouths by devising ways to eat. If the anastomosis can be kept open to some extent, it is possible to eat by mouth. In this respect, it is also desirable to find a way to reduce esophageal restenosis as much as possible. Such unmet medical needs are presumed to exist all over the world. Although this study was limited to a single case, future clinical studies should clarify in which cases cell sheet transplantation is effective and can completely prevent restenosis.

In this study, autologous cell sheets produced at a hospital equipped with a CPF were transported to another hospital for transplantation into patients without any problems in the quality and safety of the cell sheets. A 48-week follow-up examination after cell sheet transplantation also confirmed the safety of the cell sheets for the patients. If the production and distribution of cell sheets can be systematized as in this study, research on cell sheet transplantation therapy will become easier, and eventually, it could become a common treatment method. In addition, the development of a new transplantation device specifically for pediatric patients contributed greatly to the success of the transplantation in this study. Cell sheet transplantation into the esophagus of a pediatric patient was difficult to perform because the working space was much smaller than that for adults. Likewise, problems such as edema in the esophagus and bleeding from the laceration after EBD must be addressed, so proper transplantation of cell sheets requires a high level of skill. In order to address these difficulties and to make this method widely available, it will be important to increase the number of patients in our study and to develop and improve the transplantation device.

## Data Availability

The datasets used and/or analyzed during the current study are available from the corresponding author on reasonable request.

## List of abbreviations

EBD: Endoscopic balloon dilatation
ESD: Endoscopic submucosal dissection
ESCC: Esophageal squamous cell carcinoma
CPF: Cell processing facility
SOP: Standard operating procedures
DMEM: Dulbecco’s modified Eagle’s medium
EDTA: Ethylenediamine tetraacetic acid tetrasodium
KCM: Keratinocyte culture medium

## Declarations

### Ethics approval and consent to participate

All experiments with human cells and tissues were approved by the Institutional Review Board at the National Center for Child Health and Development. Human cells in this study were utilized in full compliance with the Ethical Guidelines for Medical and Health Research Involving Human Subjects (Ministry of Health, Labor, and Welfare (MHLW), Japan; Ministry of Education, Culture, Sports, Science and Technology (MEXT), Japan).

### Consent for publication

Informed consent was obtained from a parent of the patient.

### Competing interests

AU is a co-researcher with CellSeed Inc. MM is the CEO of MakeWay LLC. The other authors have no conflicts of interest regarding the work described herein.

### Funding

This research was supported by AMED; by KAKENHI; by the Grant of National Center for Child Health and Development.

### Authors’ contributions

AF, YF, and NK designed experiments. YB, NI, and TI collected tissue from the patient. RT fabricated cell sheets and performed quality control tests. AF, YF, and KA performed EBDs. YF, MA, and MM performed transplantation of cell sheets. AF and KA performed follow-up examinations. MM contributed reagents, materials, and transplantation tools. AF, YF, and AU discussed the data and manuscript. AF, YF, RT, and MM wrote this manuscript. All authors read and approved the final manuscript.

## Acknowledgements

We would like to express our sincere thanks to K. Ishikawa for providing expert technical assistance, to C. Ketcham for English editing and proofreading, to E. Suzuki for English writing, and to K. Saito for secretarial work.

## Supplemental Information

**Supplemental Table 1. Inspection items performed for epithelial cell sheet transplantation and their timing**

## Notes

### Clinical Trial

UMIN000034566

